# B-OK: A Visual and Tactile Tool for HIV Treatment Adherence Support in a United States Urban Center

**DOI:** 10.1101/2024.02.28.24303498

**Authors:** Aaron Richterman, Tamar Klaiman, Daniel Palma, Eric Ryu, Laura Schmucker, Katherine Villarin, Gabrielle Grosso, Kathleen A Brady, Harsha Thirumurthy, Alison Buttenheim

## Abstract

Lack of adherence to antiretroviral therapy (ART) and poor retention in care are significant barriers to ending HIV epidemics. Treatment adherence support (TAS) effectiveness may be constrained by limited awareness and understanding of the benefits of ART, particularly the concepts of treatment as prevention and Undetectable=Untransmittable (U=U), for which substantial knowledge gaps persist. We used mixed methods to evaluate a straightforward visual and tactile tool, the B-OK Bottles (“B-OK”), that incorporates human-centered design and behavioral economics principles and is designed to change and strengthen mental models about HIV disease progression and transmission. We enrolled 118 consenting adults living with HIV who were clients of medical case managers at one of four case management agencies in Philadelphia. All participants completed a pre-intervention survey, a B-OK intervention, and a post-intervention survey. A subset (N=52) also completed qualitative interviews before (N=20) or after (N=32) B-OK. Participants had a median age of 55 years (IQR 47-60), about two-thirds were male sex (N=77, 65%), nearly three-quarters identified as non-Hispanic Black (N=85, 72%), and almost all reported receiving ART (N=116, 98%). Exposure to B-OK was associated with improved awareness and understanding of HIV terminology, changes in attitudes about HIV treatment, and increased intention to rely on HIV treatment for transmission prevention. Insights from qualitative interviews aligned with the quantitative findings as respondents expressed a better understanding of U=U and felt that B-OK clearly explained concepts of HIV treatment and prevention. These findings provide a strong rationale to further evaluate the potential for B-OK to improve TAS for PLWH.

## Introduction

New HIV diagnoses in the United States have plateaued at around 38,000 annually since 2013.^1,2^ This lack of progress prompted the United States Department of Health and Human Services to announce a ten-year “Ending the HIV Epidemic” (EHE) initiative in 2019.^3^ The goal of EHE is to reduce new infections by 90% by 2030 through intensified diagnostic, treatment, prevention, and outbreak response efforts in the 48 counties where over half of HIV diagnoses occurred in 2016 and 2017.^3^ In many settings, including Philadelphia — one of the EHE priority jurisdictions — lack of adherence to antiretroviral therapy (ART) and poor retention in care are the most significant barriers to ending the HIV epidemic.^4^ As of 2020, among the 14,873 people living with HIV (PLWH) in Philadelphia who had been in care in the last 5 years, 14% were out of care (accounting for 36% of HIV transmissions), and 11% were in care but not virally suppressed (accounting for 25% of HIV transmissions).^3^

Various treatment adherence support (TAS) interventions administered by clinical providers, case managers, peers, and others have been shown to improve engagement in care and ART adherence.^5,6^ However, TAS effectiveness may be constrained by limited awareness and understanding of the benefits of ART, including the benefits of treatment as prevention. Despite multiple landmark studies showing that viral suppression with ART prevents HIV transmission between serodiscordant sexual partners,^7–9^ and a global “Undetectable=Untransmittable” (U=U) campaign designed to translate these findings into a movement to promote ART use,^10^ substantial knowledge gaps persist about the prevention benefits of ART and U=U remains a challenging concept to convey to individuals in a simple and scalable manner.^11,12^ In Philadelphia, knowledge barriers were reported by 79% of PLWH at re-engagement in care, with responses such as “since there is no cure for HIV, why should I go to a doctor?”^13^ Maintaining viral suppression can be an effective motivator for remaining in care and adhering to ART, but only if a PLWH understands and internalizes concepts related to HIV pathogenesis, transmission, and treatment.^12^

Behavioral economics insights, which can help explain how people process complex health information and make health-related decisions,^14^ may be instrumental in designing strategies to improve TAS. For example, behavioral economics research shows that people use simplified cognitive representations of complex concepts, referred to as mental models, during decision-making. Mental models are known to influence beliefs and behavior in many health-related domains,^15–21^ and incorrect mental models may be one reason why people do not understand what it means to be virally suppressed, or why they may underestimate the HIV prevention benefits of ART. Interventions aiming to correct mental models about HIV and ART can benefit from the use of other behavioral economics principles, such as presenting information in a salient manner, leveraging framing effects (e.g., health messages may be more effective if they avoid emphasizing disease and vulnerability), and employing analogies to introduce and reinforce new mental models.^22^

In this study, we used mixed methods to evaluate a straightforward visual and tactile tool, the B-OK Bottles (hereafter referred to as “B-OK”), that integrates these behavioral economics principles while also drawing on several components of the Extended Process Parallel Model (EPPM) for behavior change communication.^23^ B-OK was initially developed for the South African context,^24^ and in this study we aimed to explore B-OK’s potential to improve TAS for PLWH in Philadelphia. We hypothesized that this tool would correct mental models about HIV disease progression and transmission, reframe attention towards taking control of one’s health through achieving viral suppression and away from the loss of health, and enhance retention, recall, and comprehension of complex HIV-related information (Figure 1). We further hypothesized that these changes would lead to favorable changes in HIV-related awareness, knowledge, attitudes, intentions, or perceptions, ultimately increasing the likelihood of adherence, retention in care, and viral suppression.

**Figure 1.**
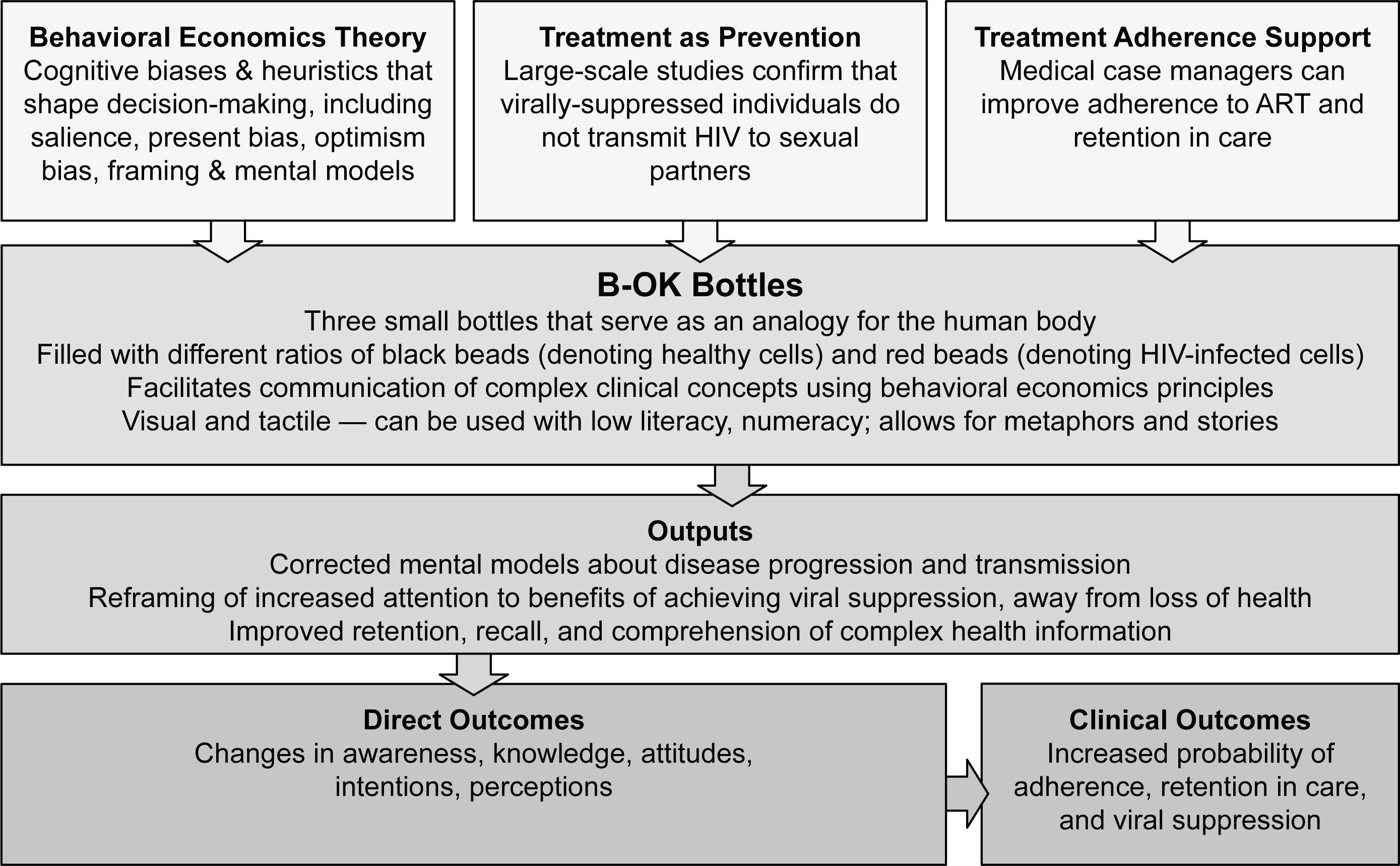
Proposed conceptual framework illustrating the hypothesized role of B-OK to facilitate treatment adherence support for people with HIV, with the goal of improving clinical outcomes.

## Methods

### Study Design and Population

We conducted a mixed methods study using an intervention (QUAN + qual) design.^25^ We enrolled PLWH who were clients of medical case managers at one of four Ryan White-funded case management agencies in Philadelphia: one community-based organization (Bebashi – Transition to Hope) and three clinic-based organizations (Drexel University, Albert Einstein Medical Center, and Temple University). Eligibility criteria required participants to be age 18 years or older and able to speak English or Spanish. Study staff recruited participants presenting for scheduled or unscheduled appointments with their medical case managers, and all study activities were completed on the same day.

All participants completed a pre-intervention survey, B-OK, and a post-intervention survey. A subset of participants also completed qualitative interviews before or after B-OK.

The University of Pennsylvania Institutional Review Board approved this study. All participants provided written informed consent. One participant withdrew from the study and was not included in this analysis. All participants received $25 compensation, and participants completing qualitative interviews received an additional $25.

### Intervention

B-OK is a non-proprietary tool designed to increase HIV treatment literacy (Figure 2). It was developed using human-centered design principles as part of Population Services International’s (PSI) Coach Mpilo project and MINA campaign in South Africa in collaboration with Matchboxology.^26^ B-OK comprises three bottles filled with colored beads in varying ratios to represent the human body in the context of HIV infection. Black beads symbolize healthy cells, while red beads represent HIV and HIV-infected cells. One bottle has a mix of black and red beads, one has mainly red beads with just a few black beads, and the third has black beads with just one red bead. The mixed bottle represents the body at HIV diagnosis, communicating that even though a PLWH might feel well and conclude that no immediate treatment is required, the presence of red beads indicates that HIV is multiplying, detectable, and transmissible. The bottle with predominantly red beads demonstrates the consequences of delaying or stopping treatment. Conversely, the bottle with only one red bead illustrates that taking ART daily can nearly (but not completely) eliminate HIV, rendering the virus undetectable, untransmittable, and virtually unable to harm a PLWH. We used two sets of bottles — a large set (each bottle measured 11 inches tall x 4.5 inches diameter) and a small set (3.75 inches x 1 inch). Both sizes were available during the intervention.

**Figure 2.**
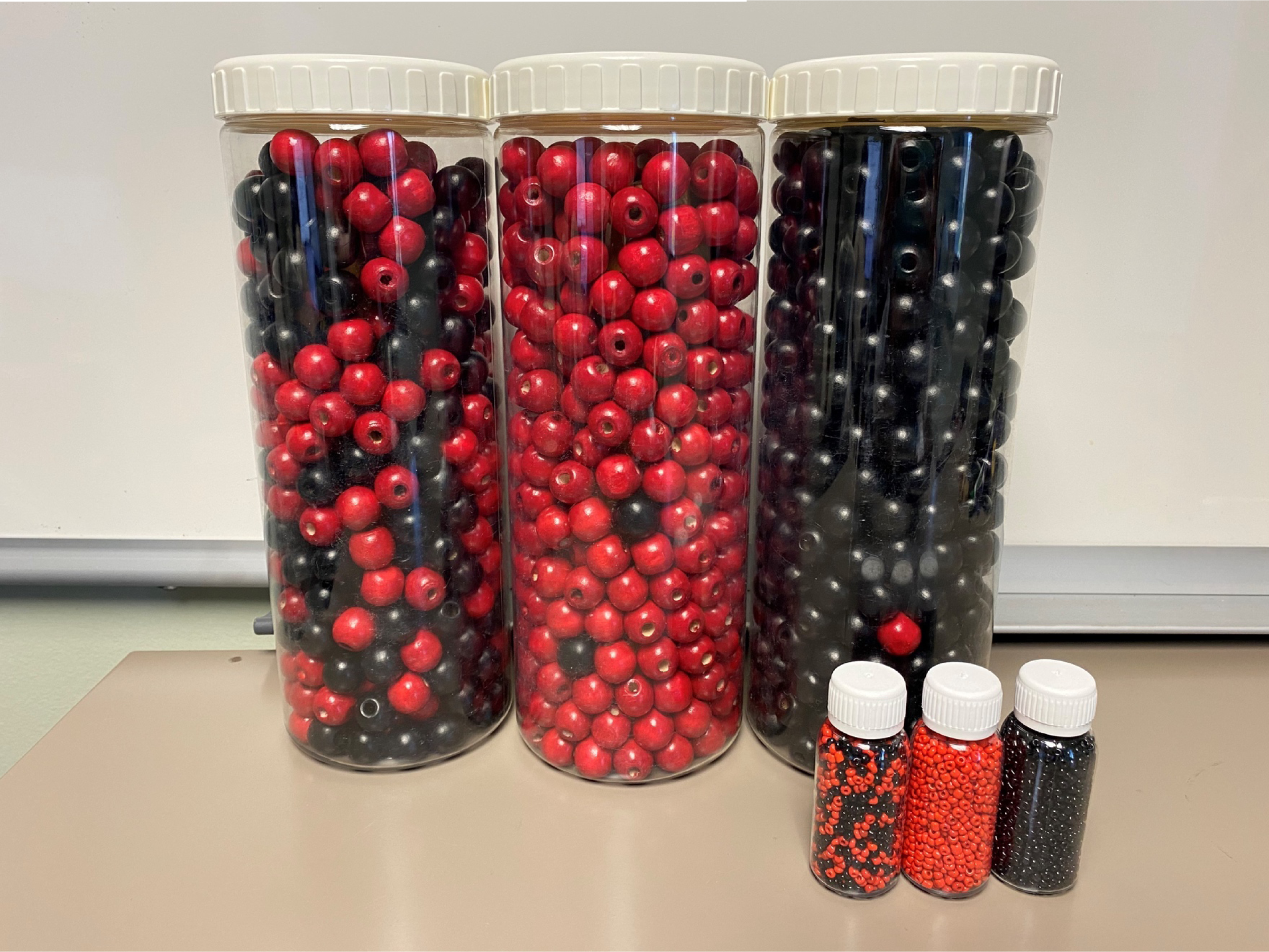
Photos of the large and small sets of B-OK Bottles.

We developed a conceptual framework illustrating the hypothesized effects of B-OK on PLWH (Figure 1). The study intervention itself was a 5-minute scripted education session administered by one of three trained study staff who were stationed at clinics or MCM offices (see Supplementary Appendix for script). The session focused on using B-OK to explain the fundamental principles of HIV biology (pathogenesis, transmission, clinical outcomes), the effects and importance of ART, and essential HIV terminology (viral suppression, undetectable viral load, and U=U). Participants were encouraged to examine or hold the bottles and to ask questions.

### Survey Questionnaires

Study staff administered pre- and post-intervention survey questionnaires to all participants. Pre-intervention surveys included questions about demographics, current ART (none, injectable, or oral), and self-reported number of missed ART doses in the last 30 days among those on oral ART (i.e., adherence). Pre- and post-intervention surveys included questions about awareness of HIV-related terminology (viral suppression, U=U), knowledge about these terms and the effects of ART on clinical outcomes and transmission risk, attitudes about treatment as prevention and prognosis on ART, intentions about relying upon treatment as prevention as a preventive strategy and intention to use ART generally, and perceptions of HIV illness understanding. Awareness questions were binary (yes or no); other questions used probabilistic scales from zero to ten (ten indicating greater knowledge, certainty, etc.).^27^ Survey questions were developed based on items used previously in formative and impact studies of U=U and treatment as prevention interventions.^12,28–37^ Surveys were conducted in English or Spanish.

### Statistical Analyses

We used descriptive statistics to summarize participant demographics and ART-related variables. Our co-primary outcomes were the 11 questions about HIV awareness, knowledge, attitudes, intentions, and perception. For each outcome, we reported summary statistics of the pre-intervention scores and changes between pre- and post-intervention scores. For the awareness variables, we compared the proportions of participants who were aware of viral suppression and U=U before and after the intervention using two-sample, two-tailed z-tests. For all other variables, we used single-sample two-tailed t-tests to assess whether the change in the score pre-to post-intervention was significantly different from zero. We reported unadjusted p-values, and p-values that were adjusted for 11 multiple comparisons using the Benjamini-Hochberg method.^38^ We considered p<0.05 to be statistically significant.

As a secondary analysis, we assessed changes in outcomes in several key subgroups: whether a participant had a pre-intervention interview (based on the hypothesis that discussion about HIV may influence survey answers); optimal adherence to oral ART (>95% over the last 30 days by self-report); and agency type (community-based or clinic-based). We compared changes in the outcomes between subgroups using two-tailed, two-sample t-tests.

Statistical analyses were conducted using Stata version 17 (StataCorp) and R version 4.2.3 (R Foundation for Statistical Computing).

### In-Depth Interviews

We conducted semi-structured qualitative interviews to explore mental models of HIV biology, treatment, and prevention. We recruited participants consecutively and interviewed them before or after exposure to B-OK. Each group (pre- and post-intervention) had a planned sample size of at least 20 interviews. We used purposive selection to ensure inclusion across the agencies. One of three study staff members trained in qualitative research (D.P., G.G., and K.V) conducted the interviews in either English or Spanish. Study staff were not employed by the case management agencies and were not involved in the participants’ care. We completed the interviews on the day of enrollment, in person, and in a private space. We audio-recorded all interviews, which were then professionally transcribed by an external company.

### Qualitative Analyses

Our codebook was developed using an integrated analysis approach that involved major themes from modified grounded theory and a priori domains related to HIV pathogenesis, treatment, and prevention.^39^ We used an iterative process to refine and finalize the codebook. Two investigators independently coded transcripts (D.P. and E.R.), double-coding 20%. Disagreements were discussed and resolved through consensus. We analyzed the data overall and after stratifying by whether the interview was pre-intervention or post-intervention. Qualitative analyses were completed using ATLAS.ti version 23 (ATLAS.ti Scientific Software Development GmbH).

## Results

### Participant Characteristics

We enrolled 118 participants with a median age of 55 years (IQR 47-60) from March to April 2023, with about half (N=55, 47%) recruited from the community-based agency (Table 1). About two-thirds were male sex (N=77, 65%), and the vast majority were cis-gender (N=113, 96%) and spoke English as their primary language (N=110, 93%). Nearly three-quarters of the participants identified as non-Hispanic Black (N=85, 72%), with the remainder identifying as Hispanic (N=19, 16%), non-Hispanic white (N=6, 5%), and other or multiple race/ethnicities (N=8, 7%). Almost all participants reported receiving ART, either oral (N=107, 91%) or injectable (N=9, 8%). Among participants receiving oral ART, the median number of reported doses missed out of the last 30 days was zero (IQR 0-2), and 36 (33%) had sub-optimal adherence (i.e., <95% self-reported adherence during the last 30 years)

**Table 1.** Characteristics of the study population. Data are presented as N (%) unless otherwise specified. N=118 unless otherwise specified.

|  |  |
| --- | --- |
| Age, median (IQR) | 55 (47-60) |
| Male sex | 77 (65) |
| Cis-gender | 113 (96) |
| English primary language (vs Spanish) | 110 (93) |
| Race / Ethnicity |  |
| Hispanic | 19 (16) |
| Non-Hispanic Black | 85 (72) |
| Non-Hispanic White | 6 (5) |
| Other/Multiple | 8 (7) |
| Site |  |
| Bebashi | 55 (47) |
| Drexel | 31 (26) |
| Temple | 25 (21) |
| Einstein | 7 (6) |
| ART |  |
| None | 2 (2) |
| Injectable | 9 (8) |
| Oral | 107 (91) |
| Number of missed oral ART doses in last 30 days, median (IQR), N=107 | 0 (0-2) |

Abbreviations: ART, antiretroviral therapy

### Changes in HIV Awareness and Knowledge, Attitudes, Intentions, and Perceptions After B-OK

Before B-OK, just over half of the participants (N=61, 52%) had ever heard of the term viral suppression, and three-quarters (N=88, 75%) recognized the term U=U. Following the intervention, awareness rose to 69% (N=81) and 86% (N=101) for viral suppression and U=U, respectively. These changes were statistically significant based on unadjusted p-values (p=0.01 for viral suppression and p=0.03 for U=U), but only the increase in viral suppression remained significant when adjusted for multiple comparisons (adjusted p=0.02 for viral suppression and adjusted p=0.06 for U=U). On ten-point scales, participants’ pre-intervention self-reported understanding about the term viral suppression had a mean of 3.4 (SD 4.1), and U=U had a mean of 6.6 (SD 4.3) (Table 2, Supplementary Figures 1-2). Both significantly increased after B-OK — by 2.4 (SD 4.0) for viral suppression (p<0.0001; adjusted p=0.001), and 1.4 (SD 3.7) for U=U (p=0.0001; adjusted p=0.0006).

**Table 2.** Responses on a 0-10 scale (with higher numbers indicating greater agreement) to questions about HIV-related knowledge, attitudes, intentions, and perceptions — baseline score, difference before and after exposure to B-OK, and p-value testing whether the difference was significantly different from zero (single sample t-test, two-tailed, both unadjusted and adjusted for 11 multiple comparisons — Benjamini-Hochberg). Data are presented as mean (SD), N=118.

|  | Baseline | Difference pre-post | p-value | Adjusted p-value |
| --- | --- | --- | --- | --- |
| **Knowledge** |  |  |  |  |
| How well do you understand the term “viral suppression?” | 3.4 (4.1) | 2.4 (4.0) | <0.0001 | 0.001 |
| How well do you understand the term “U=U” or “Undetectable=Untransmittable?” | 6.6 (4.3) | 1.4 (3.7) | 0.0001 | 0.0006 |
| Imagine a person living with HIV who is taking HIV treatment and has an undetectable viral load. How likely is it that this person would pass HIV to a regular sexual partner? | 2.1 (3.3) | 0.0 (3.6) | 0.94 | 0.94 |
| Imagine a person living with HIV who is taking HIV treatment and has an undetectable viral load. How likely is it that this person would be harmed by HIV? | 2.6 (3.6) | 0.1 (3.9) | 0.74 | 0.81 |
| **Attitudes** |  |  |  |  |
| Agrees with: I worry that HIV medications do not completely eliminate the risk of getting HIV through sex | 4.3 (4.3) | -0.8 (4.4) | 0.05 | 0.08 |
| Agrees with: Having HIV means I will eventually die from it | 3.1 (4.1) | -0.8 (3.4) | 0.008 | 0.02 |
| **Intentions** |  |  |  |  |
| Imagine you were taking HIV treatment and had an undetectable viral load. How likely would you be to use a condom to prevent passing HIV to a sex partner who did not have HIV? | 7.8 (3.3) | -1.2 (3.0) | 0.0001 | 0.0006 |
| Agrees with: I feel motivated to stay on (or restart) HIV treatment | 9.5 (1.8) | 0.1 (1.7) | 0.38 | 0.46 |
| **Perceptions** |  |  |  |  |
| Agrees with: I understand my illness | 9.2 (2.0) | 0.2 (1.6) | 0.23 | 0.32 |

Abbreviations: HIV, human immunodeficiency virus; U=U, Undetectable=Untransmittable

Participants had high pre-intervention knowledge when asked to imagine a scenario in which a PLWH has an undetectable viral load and estimate the probability that this person would transmit HIV to a sexual partner (pre-intervention mean 2.1, SD 3.3) or be harmed by HIV (pre-intervention mean 2.6, SD 3.6), with neither item significantly changing after the intervention (p=0.94, adjusted p=0.94 for HIV transmission; p=0.74, adjusted p=0.81 for HIV harm) (Supplementary Figures 3-4).

Attitudes about the effectiveness of treatment as prevention (pre-intervention mean 4.3, SD 4.3) and HIV prognosis (pre-intervention mean 3.1, SD 4.1), both significantly improved after exposure to the B-OK bottles (manifesting as decreases in worry) with mean changes of −0.8 (SD 4.4; p=0.05) and −0.8 (SD 3.4; p=0.008), respectively (Supplementary Figures 5-6). After adjustment, changes in attitudes about HIV prognosis remained significant (adjusted p=0.02), while treatment as prevention did not (p=0.08).

Before B-OK, participants reported that they would be unlikely to rely on HIV treatment as a sole preventive strategy, as indicated by their high likelihood of using a condom to prevent sexual HIV transmission even with an undetectable viral load (pre-intervention mean 7.8, SD 3.3) (Supplementary Figure 7). The mean change after the intervention was −1.2 (SD 3.0), representing a statistically significant increase in the likelihood of using HIV treatment as a primary preventive strategy (p=0.0001; adjusted p=0.0006).

Participants’ intentions to continue ART were very high at baseline (pre-intervention mean 9.5, SD 1.8), and did not significantly change after the intervention (p=0.38, adjusted p=0.46) (Supplementary Figure 8). Similarly, participants’ perceptions about understanding their illness were also high (pre-intervention mean 9.2, SD 2.0), without significant changes after the intervention (p=0.23, adjusted p=0.32) (Supplementary Figure 9).

### Sub-Group Analyses

Most changes pre-to post-intervention were similar across key sub-groups, with several notable exceptions (Supplementary Table 1). Attitudes about treatment as prevention improved more in the group with the post-intervention interview (mean change −1.2, SD 4.3) compared to those who had a pre-intervention interview (mean change 1.3, SD 4.6) (p=0.02), and among participants seen at the community-based site (mean change −1.9, SD 4.7) compared to the clinic-based site (mean change 0.1, SD 4.0) (p=0.02). Perceptions about illness understanding improved more among participants with sub-optimal self-reported adherence (mean change 0.6, SD 1.8) compared to those with optimal self-reported adherence (mean change −0.1, SD 1.5) (p=0.03).

### Qualitative Interviews

Fifty-two participants (44%) completed in-depth interviews — 20 (17%) before the intervention and 32 (27%) after the intervention.

Conceptions of HIV Transmission and Pathogenesis

Most respondents knew that untreated HIV would make them ill; however, others were unsure about how HIV affects the body.

> I: And what happens once the virus starts taking over those cells?
>
> R: You get AIDS, and you become susceptible to all kinds of diseases and everything like that. Your immune system shut down. (Participant 41 – post-intervention)
>
> Well, your white cells deplete, and the HIV starts to take over, killing the white T cells. And eventually it will overcome you and you’ll get sick and die. (Participant 19 – pre-intervention)
>
> What do I think happens inside the body if you have HIV? I’m not exactly sure. What do I think about – sometimes – okay, sometimes I think when you get HIV in your body I think about death, that you’re about to die was never taught about the HIV, so that’s why – as me finding out what I had, I thought I was going to die right then and there. (Participant 5 – post-intervention)
>
> Most respondents from both interview groups knew that HIV is spread through sex or exposure to bodily fluids.
>
> Because my viral load, it’s in my sperm. It’s in all the fluids in my body, my blood, my sperm. So passing it on to someone else without any protection for that person, like using a – sharing a needle, intravenous needle, or having unprotected sex or open wounds. You know what I mean? (Participant 6 – post-intervention)

Viral Load Suppression

> Results from the interviews indicated that many respondents in both groups had some understanding of viral suppression.
>
> Your viral load is very important because your viral load is the one that tell you if you undetectable or not. (Participant 9 – post-intervention)
>
> …with the treatment, it counts what your viral load is. So if you’re on your medicine or whatever treatment that you have to be on, then the more you take your medicine, the less of your viral load. The less viral load you have, to count. (Participant 20 – post-intervention)
>
> Beings though they’re taking it regularly, it suppresses a lot of the HIV. Basically, till you’re almost – that you have none, really, measurable. (Participant 19 – pre-intervention)

Others expressed confusion about the concept of viral suppression.

U=U

> I: What does the concept, viral load, mean to you?
>
> R: The viral load?
>
> I: Yes.
>
> R: I’m not sure in it. Yeah, I’m not sure if I ever knew.
>
> I: That’s no problem.
>
> R: I’m not sure. Neither one, the CD – the CD, what’s that? It’s the viral load and what else? It’s – oh God. (Participant 1 – post-intervention)
>
> Viral load is the blood cells, the white – or I forget which one. It’s the white or red. But one has to be high, and one has to be low. I’m not sure which one it is. Right? (Participant 37 – pre-intervention)
>
> Most respondents were aware that consistent treatment adherence eliminates the risk of sexual transmission (U=U).
>
> Okay. If I’m passing it, you know, if you take your medication every day, you keep your viral load low, you can’t pass the disease on to nobody. Because you’re undetectable, there’s no amount of disease present which causes you to pass on to another person. (Participant 2 – post-intervention)
>
> And with the HIV drugs, it kills those new cells or whatever. So the healthy ones can build themselves back up, and once they get back up then you are U=U. You can’t pass it on to somebody else. (Participant 11 – post-intervention)

However, several respondents expressed uncertainty about U=U —that they thought (or worried) that it was still possible to transmit HIV even with an undetectable viral load.

> Because I be saying to myself, well, I’m non-detected, I can’t pass it on. But I don’t think that’s true, because as long as it’s in my body and I do that sex thing with the blood and that blood-to-blood transfer, I think I could pass it. I believe I can. (Participant 21 – post-intervention)

The B-OK Bottles

As shown in the quantitative results, respondents reported improved understanding of viral load and the impact of not taking medication after exposure to B-OK.

> The viral load is what keeps our immune system strong or weak. When we have a very high viral load, it’s because our immune system is weakened. That is what the red bottle is for. We have a high viral load. And our body is practically defenseless. We have to take the medication to lower the viral load until it’s completely neutralized. (Participant 28 – post-intervention)
>
> I learned that – how the HIV is in reference to the first bottle which is a mixture of the dark and the red balls. And the other bottle, which is all red, which meaning that’s completely all with HIV up in there, when the dark one means that’s the best to have, try to get. (Participant 3 – post-intervention)
>
> Well, I didn’t know. Since I never seen the cells before, I didn’t know that if you’re HIV, it’s not good. All the cells are being red. But I’m here with the black bottles. I don’t want to get in between. (Participant 53 – post-intervention)

Respondents felt that the bottle demonstration clearly explained HIV and what would happen if patients stopped taking their medication or took it regularly.

> Well, it would definitely be very helpful for when [providers] explain about the viral load or about just having HIV and it entering your body, and what it would do if you take your meds, opposed to not. Well, take treatment, opposed to not taking treatment. And I mean, it’s a pretty great idea so they – people can understand at all levels. (Participant 50 – post-intervention)

## Discussion

In this mixed methods study of 118 PLWH who were clients of medical case managers in Philadelphia, we found that exposure to B-OK was associated with improved awareness and understanding of HIV terminology, changes in attitudes about HIV treatment, and increased intention to use HIV treatment as prevention. Insights from qualitative interviews aligned with the quantitative results as respondents expressed a better understanding of U=U and felt that B-OK clearly explained concepts of HIV treatment and prevention.

These findings provide evidence supporting the potential for B-OK to improve TAS for PLWH, especially considering the known persistent knowledge gaps about the prevention benefits of ART.^11–13^ This potential is further supported by prior randomized controlled trials that have demonstrated that behavioral interventions designed in part to improve understanding of treatment as prevention among PLWH can improve ART adherence, viral suppression, and intentions to rely on treatment as prevention.^36,37,40^

There were survey questions within some domains that did not improve after exposure to B-OK — in particular, hypothetical scenarios asking participants to estimate the likelihood of sexual transmission from an undetectable partner and the likelihood of harm from HIV for a person with an undetectable viral load, intentions to stay on HIV treatment, and perceived understanding of illness. Participants had very high baseline scores for these questions, so it is unsurprising that there were no significant changes seen after the intervention.

Our study population reflected the demographics and racial composition of PLWH in Philadelphia.^41^ While we did not collect detailed socioeconomic data, all participants were seen at Ryan White-funded case management sites and therefore met Ryan White eligibility by having income less than 500% of the federal poverty level. Participants generally had long-standing HIV, and had a median age of 55. Our study population was likely to have had adherence challenges at some point based on their engagement with MCMs, and one-third reported current sub-optimal adherence. It is likely that the impact of B-OK could be even greater among younger individuals with more recent (or newly diagnosed) HIV infection and less treatment experience as well as PLWH who have dropped out of care and are not engaging with MCMs. Similarly, it’s conceptually plausible that B-OK would be even more effective among PLWH with lower rates of adherence, among PLWH experiencing virological failure, or in populations with low health literacy. These are also the populations with the greatest need for TAS. This hypothesis is supported by our finding of greater improvements in self-reported illness perception among participants with sub-optimal self-reported adherence.

It is notable that the distributions of most survey responses were highly bimodal, with responses clustered at the extremes and generally a higher peak at the more desired extreme (high self-rated knowledge, greater intention to use HIV treatment as prevention, etc.). This suggests that B-OK may be particularly relevant for the relatively smaller subset of PLWH with incorrect mental models of HIV (i.e., with responses on the less desired extreme), supporting its use as part of a package of interventions for TAS, each targeting different mechanisms of non-adherence.

Our study has several important limitations. This was a study designed to assess for preliminary evidence of B-OK effectiveness, and therefore we focused on a population of all-comers at MCM agencies over a short period of time. While these results are promising, it will be important in to evaluate in populations with higher rates of non-adherence in future work, with longer term outcomes. It remains unknown whether the changes associated with B-OK identified in this study persist beyond the short-term, and whether these changes would be further reinforced by longitudinal engagement with the bottles. Similarly, we do not yet know whether there will be improvements in clinical outcomes of patients due to any changes in knowledge, attitudes and intentions that occur after exposure to B-OK. These questions should be addressed in future research. Despite this, there is now strong justification to further evaluate B-OK, either alone or as part of a package of interventions, to enhance TAS for PLWH. As part of this study, we also collected information regarding B-OK acceptability, feasibility, and appropriateness from both clients and medical case managers — these findings will be presented separately.

### Conclusion

In this mixed methods study of 118 PLWH who were clients of medical case managers in Philadelphia, we found that exposure to B-OK was associated with improved awareness and understanding of HIV terminology, changes in attitudes about HIV treatment, and increased intention to rely on HIV treatment as prevention as a primary prevention strategy. Qualitative interviews aligned clearly with the survey results as respondents expressed a better understanding of U=U and felt that B-OK clearly explained concepts of HIV treatment and prevention. Our findings provide a strong justification to further evaluate B-OK, either alone or as part of a package of interventions, to improve TAS for PLWH.

## Declarations

### Ethics approval and consent to participate

The study was approved by the Institutional Review Board of the University of Pennsylvania. Written informed consent was obtained from all participants. One participant withdrew from the study and was not included in this analysis.

## Availability of data and material

Data are available upon reasonable request to the senior author.

## Author’s Contributions

AR, HT, and AB conceived of the study. RC, DP, KV, and GG conducted data collection under supervision of AR, LS, and AB. AR conducted quantitative analyses with input from HT and AB. ER and DP conducted qualitative analyses under supervision of TK. AR wrote the first draft of the manuscript, with critical input from all authors.

## Data Availability

Prior to publication, we will make a de-identified dataset available in a public repository that is sufficient to replicate all study findings.

## Acknowledgments

We acknowledge Letitia Rambally Greener, Shawn Malone, and Population Services International, as well as Cal Bruns and Matchboxology for developing and disseminating B-OK. We are grateful to our primary contacts at each of the partner sites for their logistical assistance in completing this work: Terence Carroll and Dr. Amy Althoff at Drexel University, Dr. Ellen Tedaldi at Temple University, Aviva Joffe at Einstein Medical Center, and Nafisah Houston at Bebashi — Transition to Hope.

## Supplementary Appendix

### Intervention Guide

Now I want to show you the B-OK bottles. Please feel free to look at or hold the bottles while I talk. These bottles are to help you understand what is happening inside your body.

*Mix of black and red beads*

- First, let’s look at this bottle.
- It has a mix of black and red beads.
- The black beads show healthy cells in your body.
- The red beads show the HIV virus.
- This is what it looks like when most people first find out they have HIV, before starting treatment.
- They may feel completely normal, but the red beads show the virus is growing.
- If folks are having sex, they can pass HIV to their partners.

*Mostly red beads*

- If a person does not start HIV treatment, eventually their body will look like this red bottle.
- There are very few healthy black beads left.
- This might happen slow, or it might happen quickly, but it will happen eventually.
- At this stage, it is hard to fight off other sicknesses like cancer and infections.
- Folks often end up in the hospital and can even die.

*Mostly black beads*

- On the other hand, if a person starts HIV treatment, and takes their treatment every day, their body quickly becomes like this bottle.
- It is full of healthy black beads.
- This is what people mean when they talk about an “undetectable viral load” or a “suppressed viral load.”
- You can see there is still one red bead, because there is not yet a cure for HIV.
- However, it is so small and deeply buried that it can’t hurt you and it can’t be passed on through sex, as long as you keep taking your HIV treatment.
- This is what people mean when they say “Undetectable=Untransmittable” or “U=U”

— that if you are in this stage, you can’t pass the virus through sex, even if you don’t use a condom.

---

**Supplementary Figure 1.**
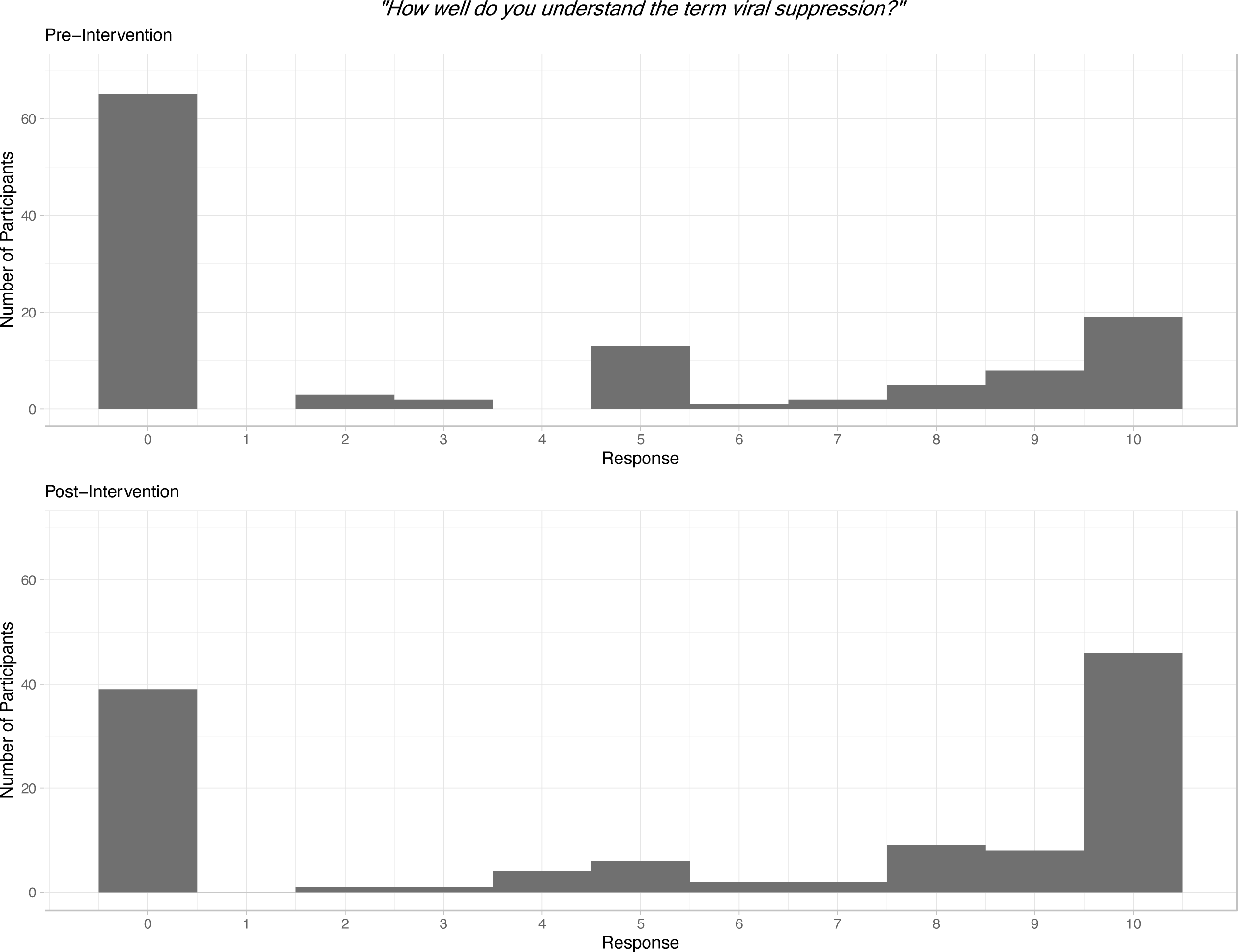
Histogram showing pre- and post-intervention responses on a zero to ten scale to the question: “How well do you understand the term ‘viral suppression?’”

**Supplementary Figure 2.**
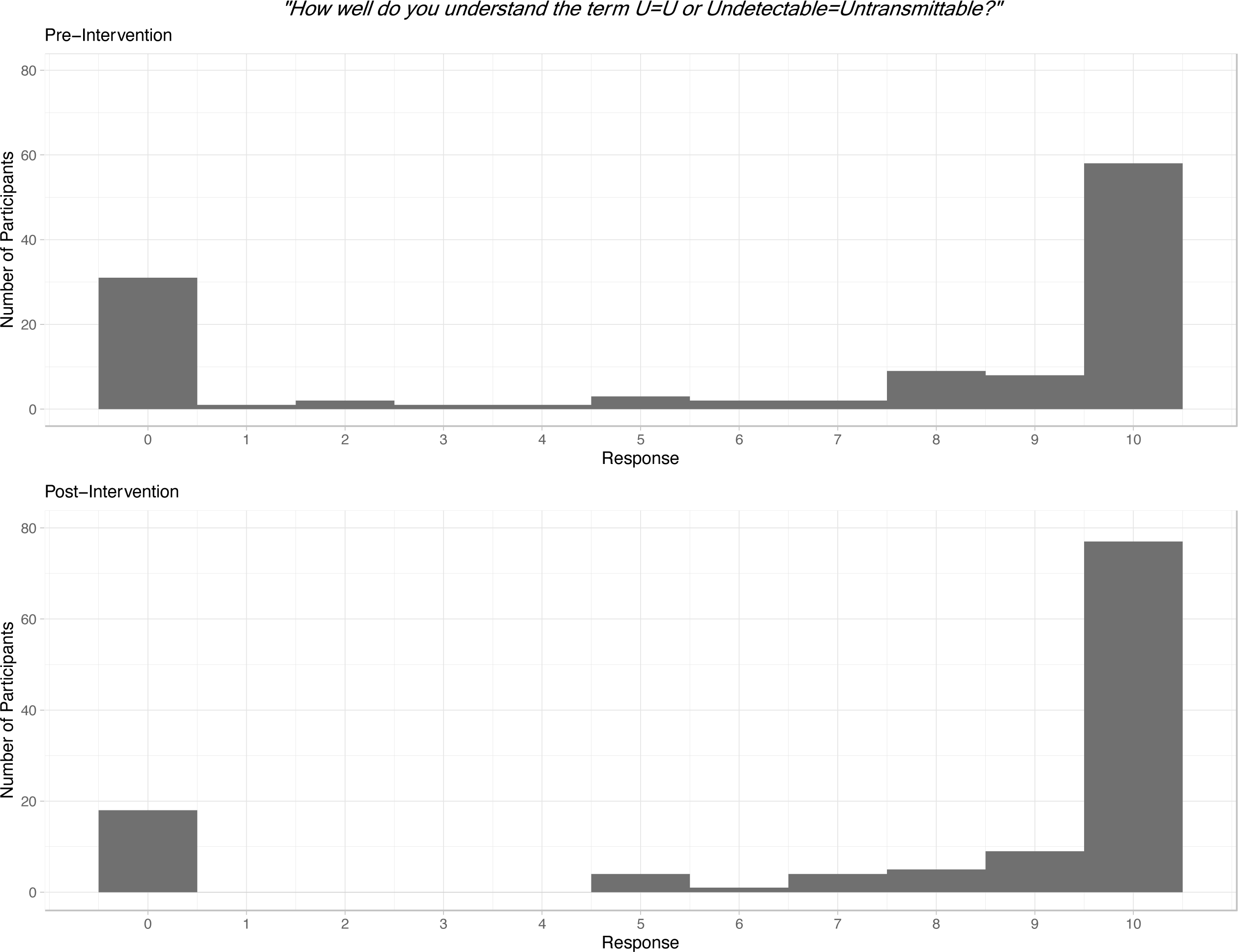
Histogram showing pre- and post-intervention responses on a zero to ten scale to the question: “How well do you understand the term ‘U=U or Undetectable=Untransmittable?’”

**Supplementary Figure 3.**
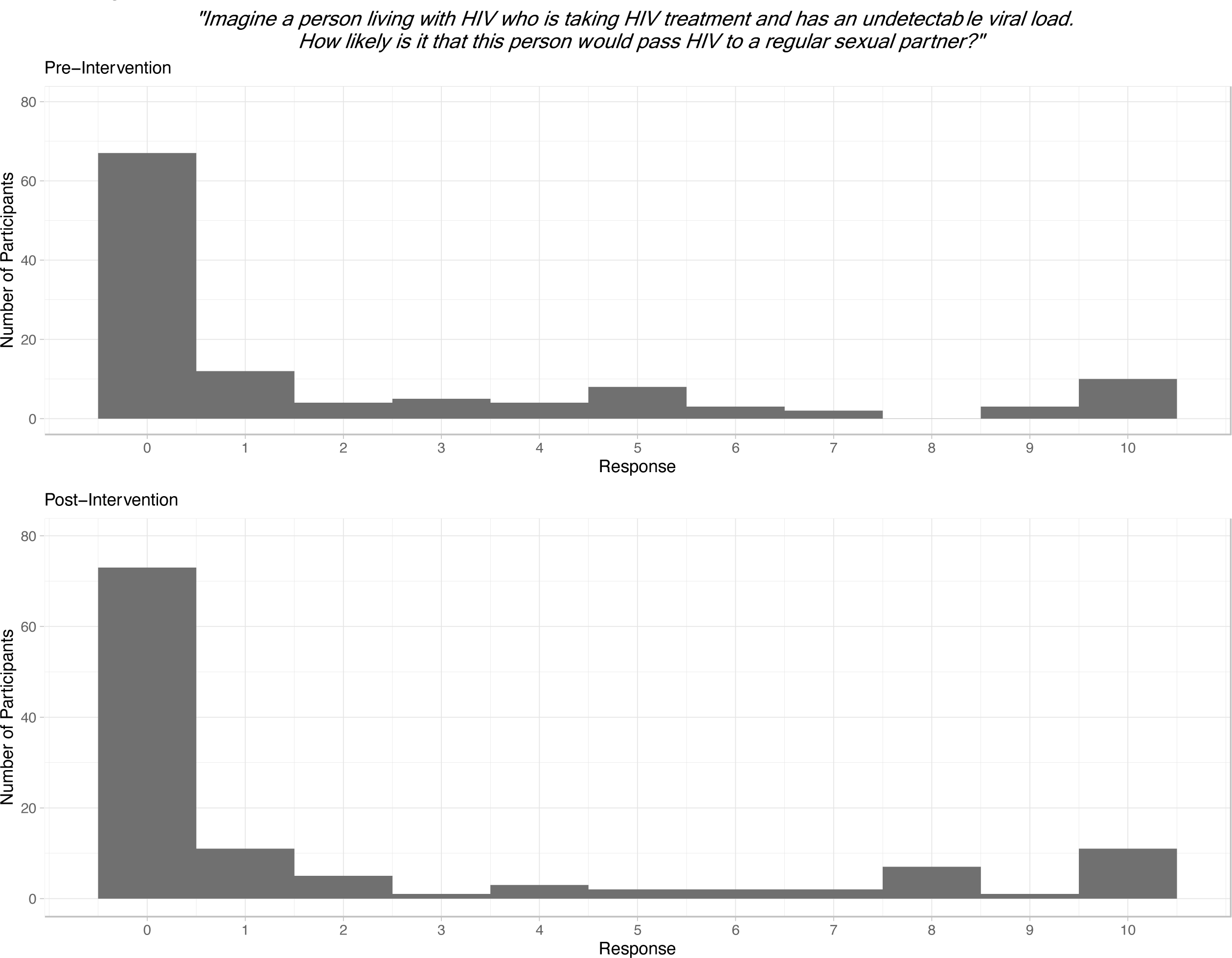
Histogram showing pre- and post-intervention responses on a zero to ten scale to the question: “Imagine a person living with HIV who is taking HIV treatment and has an undetectable viral load. How likely is it that this person would pass HIV to a regular sexual partner?”

**Supplementary Figure 4.**
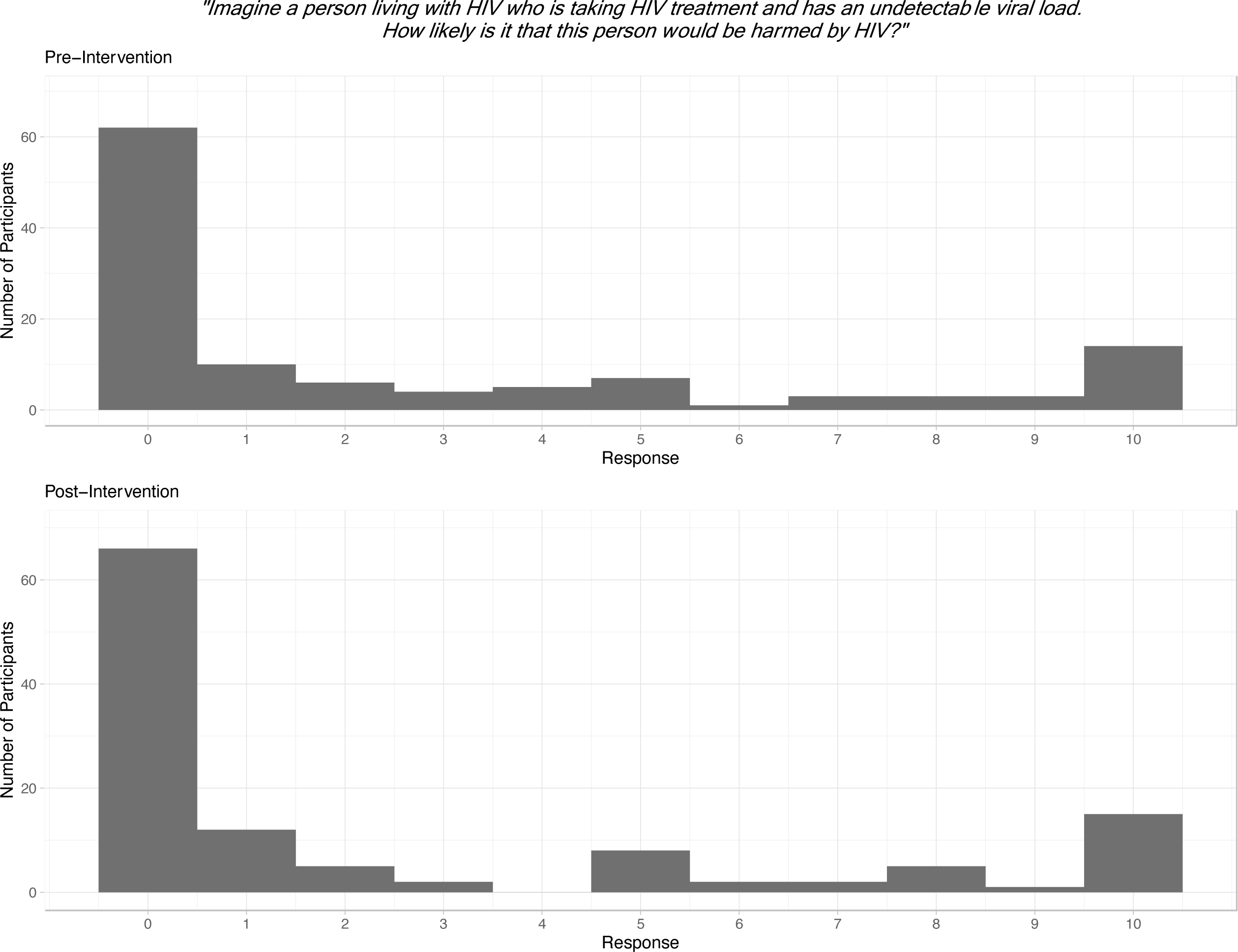
Histogram showing pre- and post-intervention responses on a zero to ten scale to the question: “Imagine a person living with HIV who is taking HIV treatment and has an undetectable viral load. How likely is it that this person would be harmed by HIV?”

**Supplementary Figure 5.**
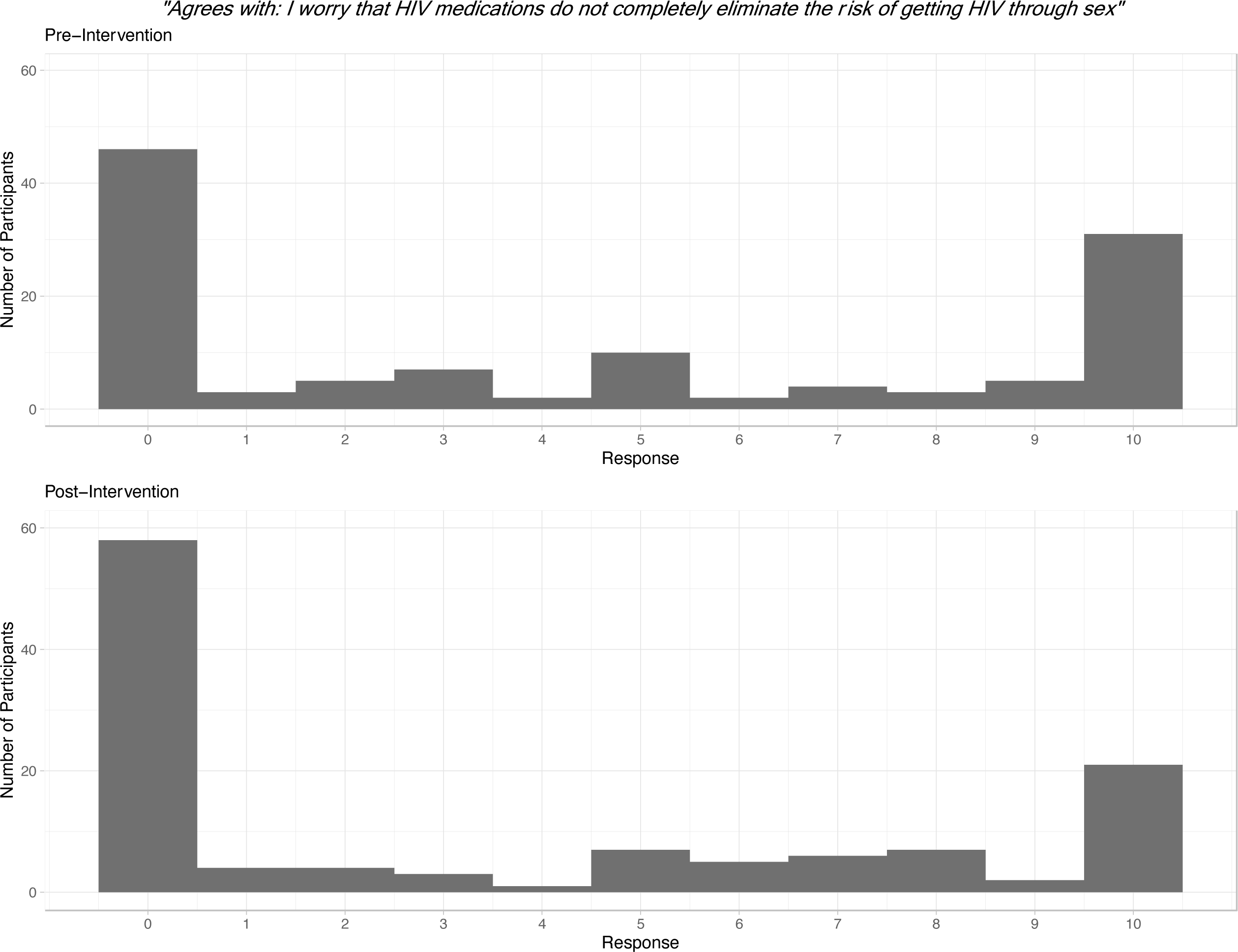
Histogram showing pre- and post-intervention responses on a zero to ten scale to the question: “Agrees with: I worry that HIV medications do not completely eliminate the risk of getting HIV through sex.”

**Supplementary Figure 6.**
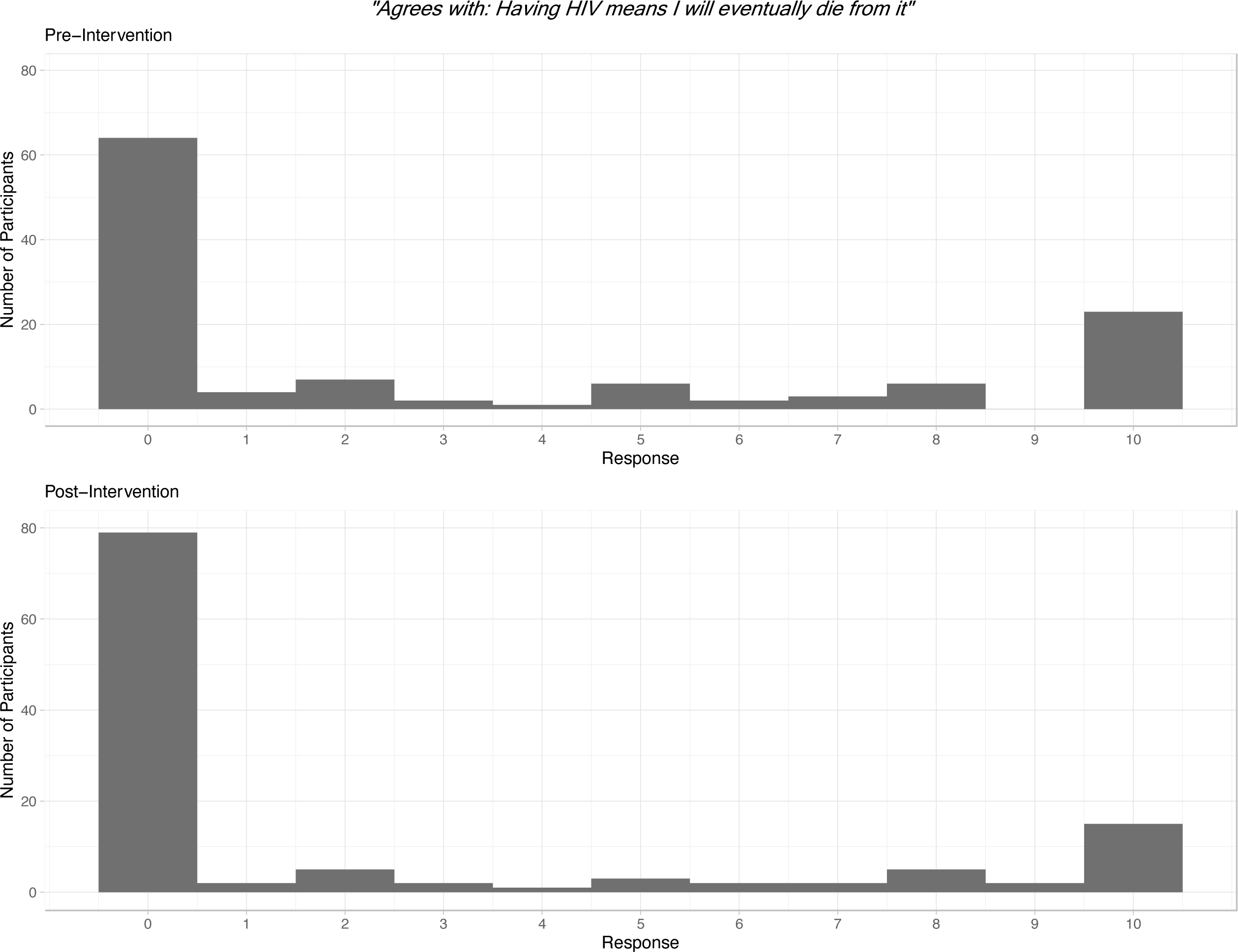
Histogram showing pre- and post-intervention responses on a zero to ten scale to the question: “Agrees with: Having HIV means I will eventually die from it.”

**Supplementary Figure 7.**
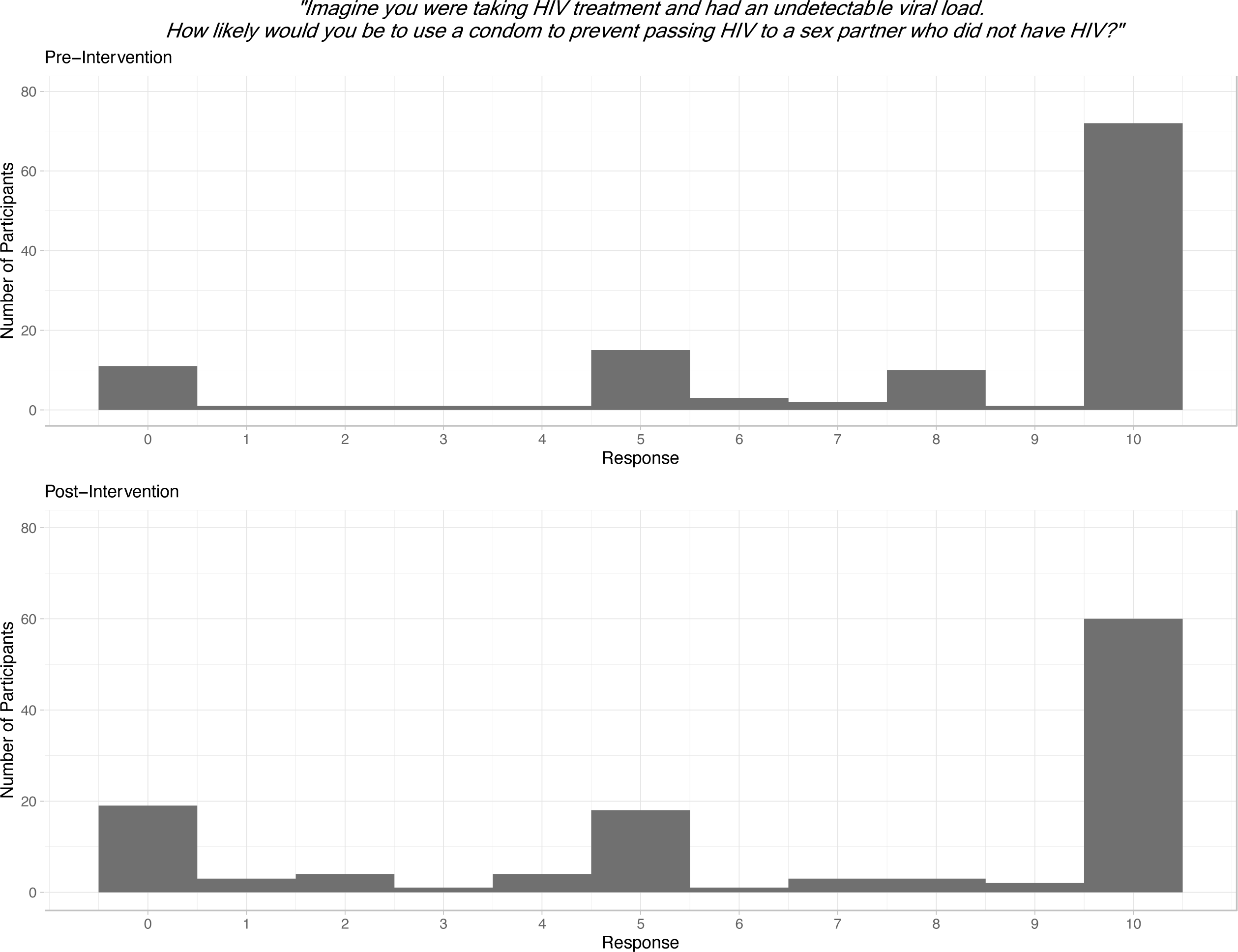
Histogram showing pre- and post-intervention responses on a zero to ten scale to the question: “Imagine you were taking HIV treatment and had an undetectable viral load. How likely would you be to use a condom to prevent passing HIV to a sex partner who did not have HIV?”

**Supplementary Figure 8.**
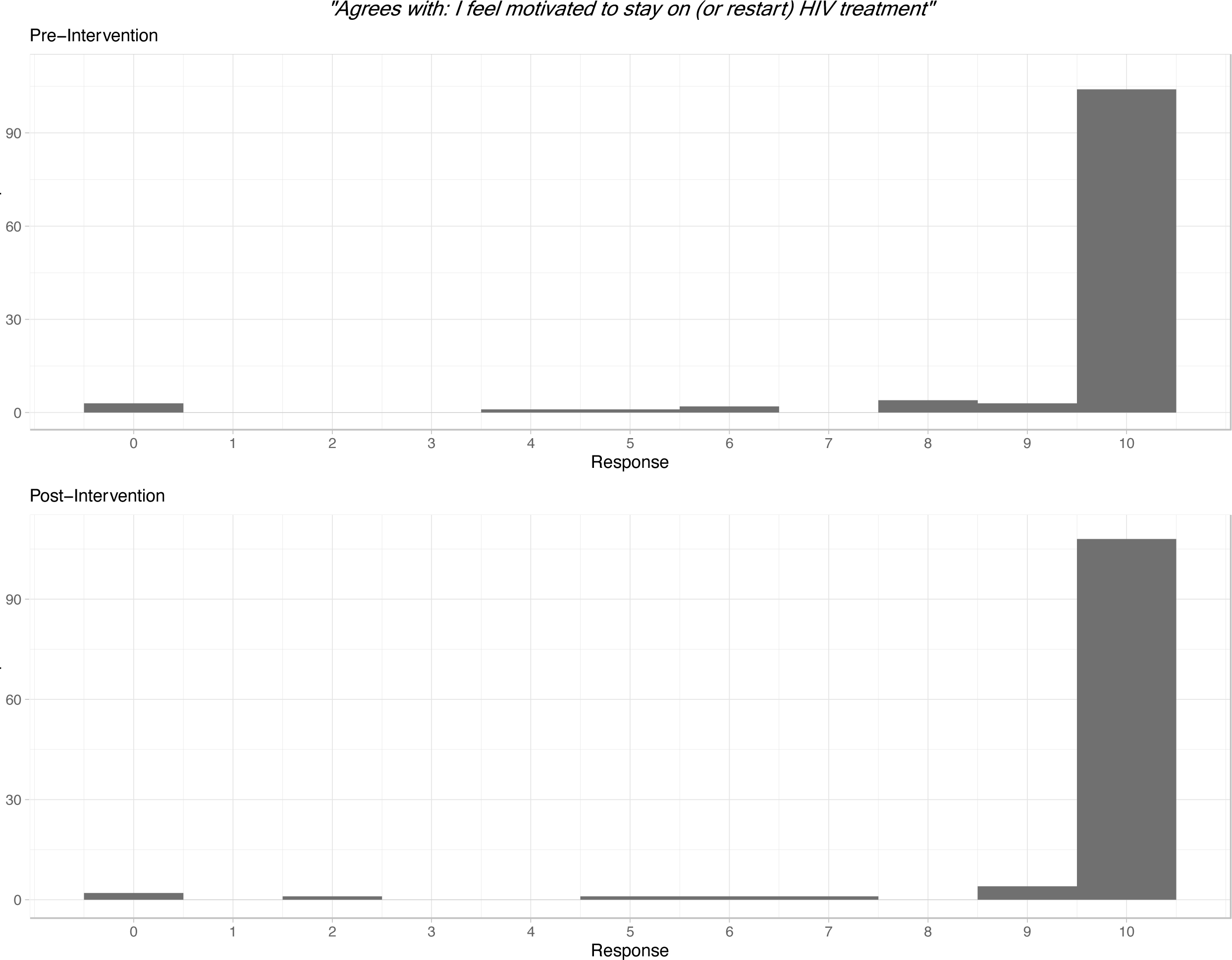
Histogram showing pre- and post-intervention responses on a zero to ten scale to the question: “Agrees with: I feel motivated to stay on (or restart) HIV treatment.”

**Supplementary Figure 9.**
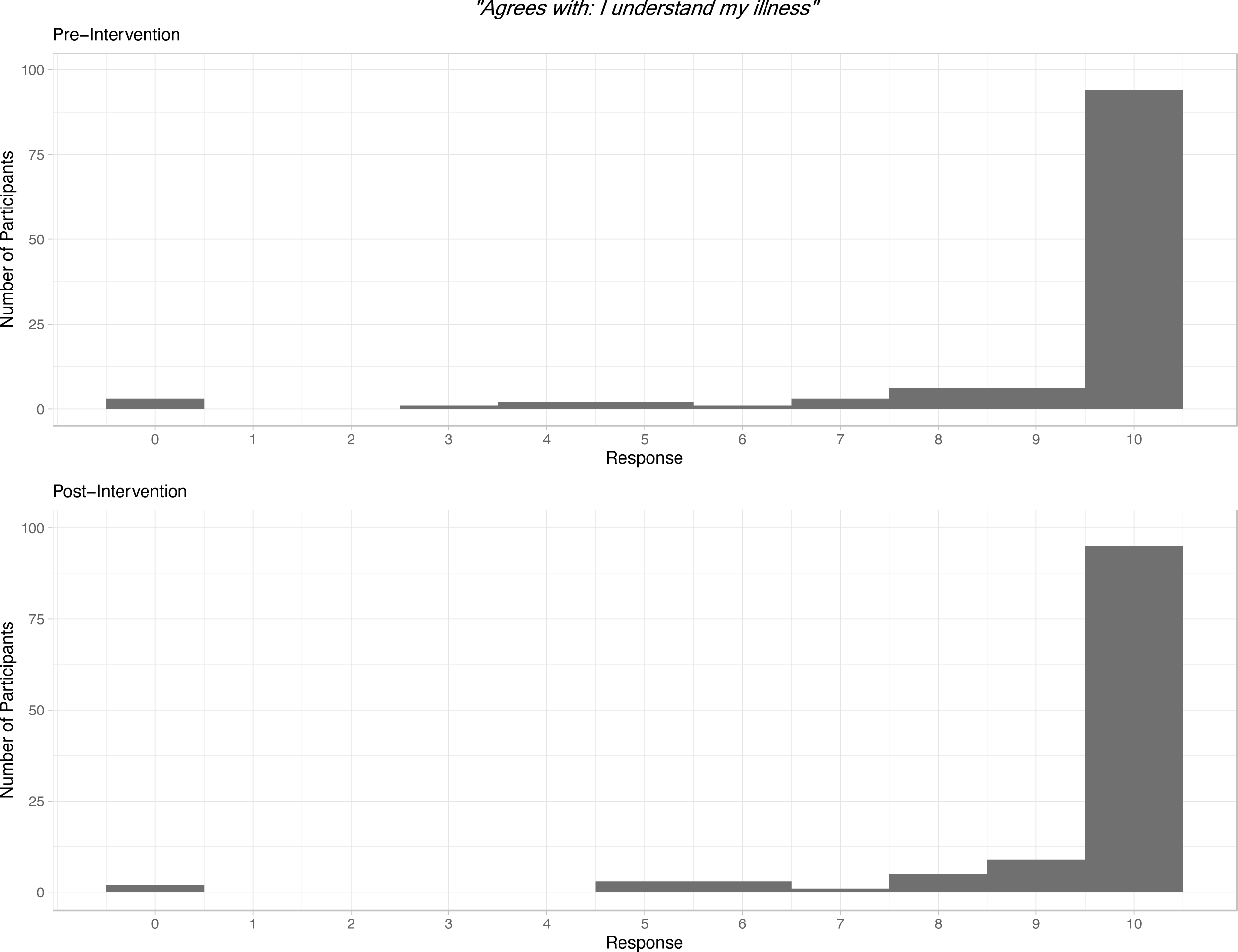
Histogram showing pre- and post-intervention responses on a zero to ten scale to the question: “Agrees with: I understand my illness.”

**Supplementary Table 1.**
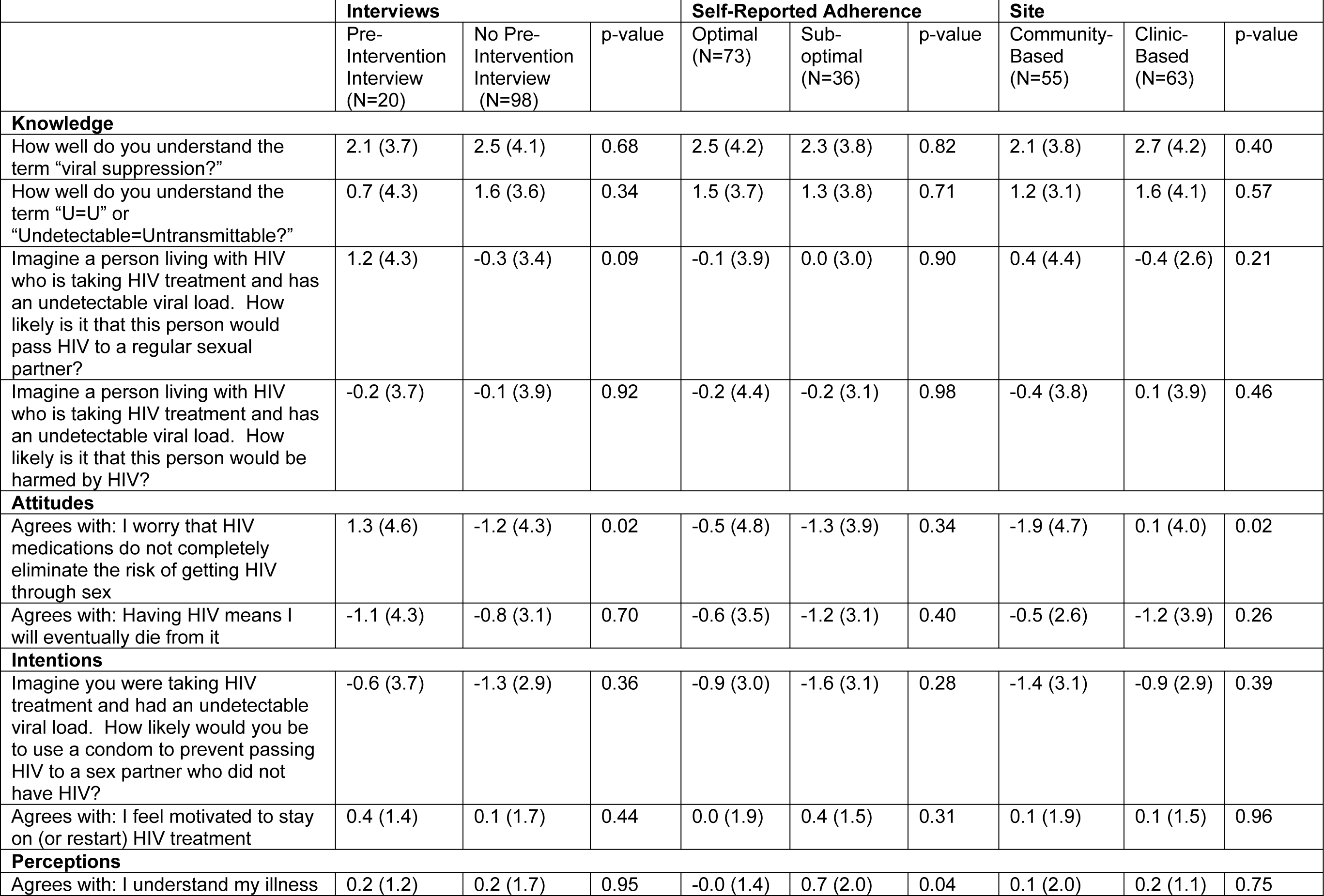
Changes in HIV-related knowledge, attitudes, intentions, and perceptions (with higher numbers indicating greater agreement to the questions) by whether participants were interviewed prior to exposure to B-OK, whether participants had optimal (>95%) self-reported adherence, and study site (community-based organization [CBO] vs university). Data are presented as mean (SD), with p-value testing whether changes were different by group (two-sample t-test, two-tailed).

